# Trio-Based Whole-Genome Sequencing for Critically Ill Pediatric Patients in Korea

**DOI:** 10.1101/2025.01.16.25320442

**Authors:** Seungbok Lee, June-Young Koh, Joonoh Lim, Jaeso Cho, Woojoong Kim, Yuna Lee, Boram Yi, Eunjung Joo, Dawoon Jung, Byung Chan Lim, Soo Yeon Kim, Jong-Hee Chae

## Abstract

This study aimed to implement genome sequencing using an automated pipeline for critically ill pediatric patients within a real-world healthcare system. Twenty patients under 36 months of age, admitted to the NICU/PICU or suspected of having rapidly progressive genetic disorders, were enrolled. Trio-based genome sequencing was performed using an optimized processing pipeline, which automatically performed mapping, variant calling, annotation, and in silico pathogenicity assessment. Among 20 enrolled patients, 11 (55%) were from the NICU, and 16 (80%) presented with neurological manifestations as their chief complaint. The median time from symptom onset to study enrollment was 73 days for 18 patients referred from other hospitals and less than a week for 2 in-hospital patients. The median turnaround time (TAT) for rWGS was 10 days, with the shortest being 5 days. A definite or presumed genetic diagnosis was made in 11 patients (55%), including 10 of 16 with neurological symptoms (62.5%) and 1 of 4 with non-neurologic symptoms (25%). Management plans were modified for 8 of the 11 patients (72.7%), including medication changes, diet modifications, and preimplantation genetic testing for future pregnancies. This study highlights the feasibility and clinical utility of WGS in critically ill pediatric patients in Korea, demonstrating a high diagnostic yield and significant impact on patient management, particularly among those presenting with neurologic symptoms. Establishing a nationwide fast-track system and providing detailed testing indications are required for effective implementation. Further automation and resource optimization could reduce the TAT and improve the efficacy of rWGS in critical care settings.

## Introduction

Since the development of next-generation sequencing, extensive research has been conducted on its clinical application, leading to its widespread use in clinical practice. Driven by progress in genomic technology, recent studies have increasingly focused on implementing whole-genome sequencing (WGS) as a first-tier approach, moving beyond gene panels and whole-exome sequencing. ^1–3^

There is a growing trend in research focusing on the use of rapid WGS (rWGS) in patients requiring urgent medical decisions. Rapid genetic testing and prompt decision making have the potential to improve clinical outcomes. We previously developed a rapid gene panel sequencing platform targeting clinically actionable genes and reported its clinical utility. ^4^ However, owing to the inherent limitations of a targeted approach that primarily screens for specific genes and variant types, there may be patients for whom a diagnosis cannot be reached.

The concept of rWGS, which enables fast-track genome-wide screening, was first proposed more than a decade ago. ^5^ Since then, it has been mostly applied to patients in neonatal intensive care units (NICU) or pediatric intensive care units (PICU). ^6–8^ However, recent studies have further demonstrated the feasibility and efficacy of rWGS in specific types of medical conditions, such as epilepsy and congenital heart disease. ^9, 10^ Technological advancements have led to the introduction of ultra-rapid long-read WGS, which demonstrates that the process from sample preparation to variant detection can be completed within 8 hours. ^11^

Considering these technological advancements and cost reductions in the field of genomic medicine, it is necessary to prepare for the utilization of rWGS in clinical settings. Although the feasibility of rWGS to deliver results within a few days has been demonstrated in previous studies, its actual implementation in healthcare systems across different societies may present an additional challenge, depending on the availability of resources and healthcare delivery system. In particular, most patients cannot undergo immediate rWGS after symptom onset because only a limited number of hospitals have the capacity to perform this procedure. This must also be taken into consideration. Here, we conducted a proof-of-concept study of rWGS targeting critically ill patients or those requiring urgent medical decisions, and examined its utility, limitations, and areas for improvement within the context of the Korean healthcare system.

## Materials and Methods

### Study cohort

The study protocol was approved by the Institutional Review Board of Seoul National University Hospital (approval no. 2303-168-1417), and compliance with the relevant guidelines and regulations was ensured. Informed consent was obtained from the parents or legal guardians of all participants prior to blood sample collection.

The prospective validation in clinical setting was conducted between March 2023 and June 2024 at Seoul National University Hospital. Patients <36 months of age were enrolled based on the following criteria: (i) admission to the NICU or PICU without acquired conditions or known genetic etiologies, or (ii) suspicion of rapidly progressive genetic disorders such as metabolic diseases.

### Library construction for whole-genome sequencing

Peripheral blood samples were collected from the probands and their family members for genomic DNA extraction. The entire workflow, from genome sequencing to analysis and interpretation, was conducted using the RareVision system (Inocras Inc., San Diego, CA, USA). Genomic DNA was isolated from the blood samples using Allprep DNA/RNA kits (Qiagen, Venlo, Netherlands). DNA libraries were prepared using TruSeq DNA PCR-Free Library Prep Kits (Illumina, San Diego, CA, USA) and sequenced on an Illumina NovaSeq6000 platform, achieving an average coverage depth of 30×.

### Processing of whole-genome sequencing data

The obtained genome sequences were aligned with the human reference genome (GRCh38) using the BWA-MEM algorithm. PCR duplicates were removed using SAMBLASTER software. ^12^ Initial variant calling for sequence variants, such as single nucleotide variant (SNV) and short insertion/deletion (indel), was performed using HaplotypeCaller and Strelka2.^13,14^ Structural variations (SV) and transposable elements (TEs) were identified using Manta and MELT, ^15, 16^ respectively. For copy number variants (CNVs), the copy numbers of all known genes were inferred from the depth of the reads covering the genes and classified as either copy gain or loss.

### Automated whole-genome sequencing pipeline

We implemented a clinical-grade automated WGS analysis pipeline powered by RareVision™, which effectively offers rare disease diagnostics by WGS and bioinformatics, from sequencing data to analysis report (Figure. 1A). The automated algorithm covered read alignment, variant calling, variant annotation, variant filtering, and advanced analysis, such as assessment of pathogenicity and relevance to the phenotype and family analysis. Following variant filtering and the assessment of Mendelian inheritance patterns, de novo mutations were detected, and their potential effects were predicted. For all variant types—SNV, INDEL, SV, and TE—those with a population allele frequency (AF) greater than 5% were excluded, in accordance with the ACMG guideline’s stand-alone criterion for identifying benign (B) or likely benign (LB) mutations. ^17^ To improve pathogenicity prediction, the WGS analysis pipeline utilized an in-house algorithm that automatically integrates updated databases, enabling more accurate analysis.

**Figure. 1.**
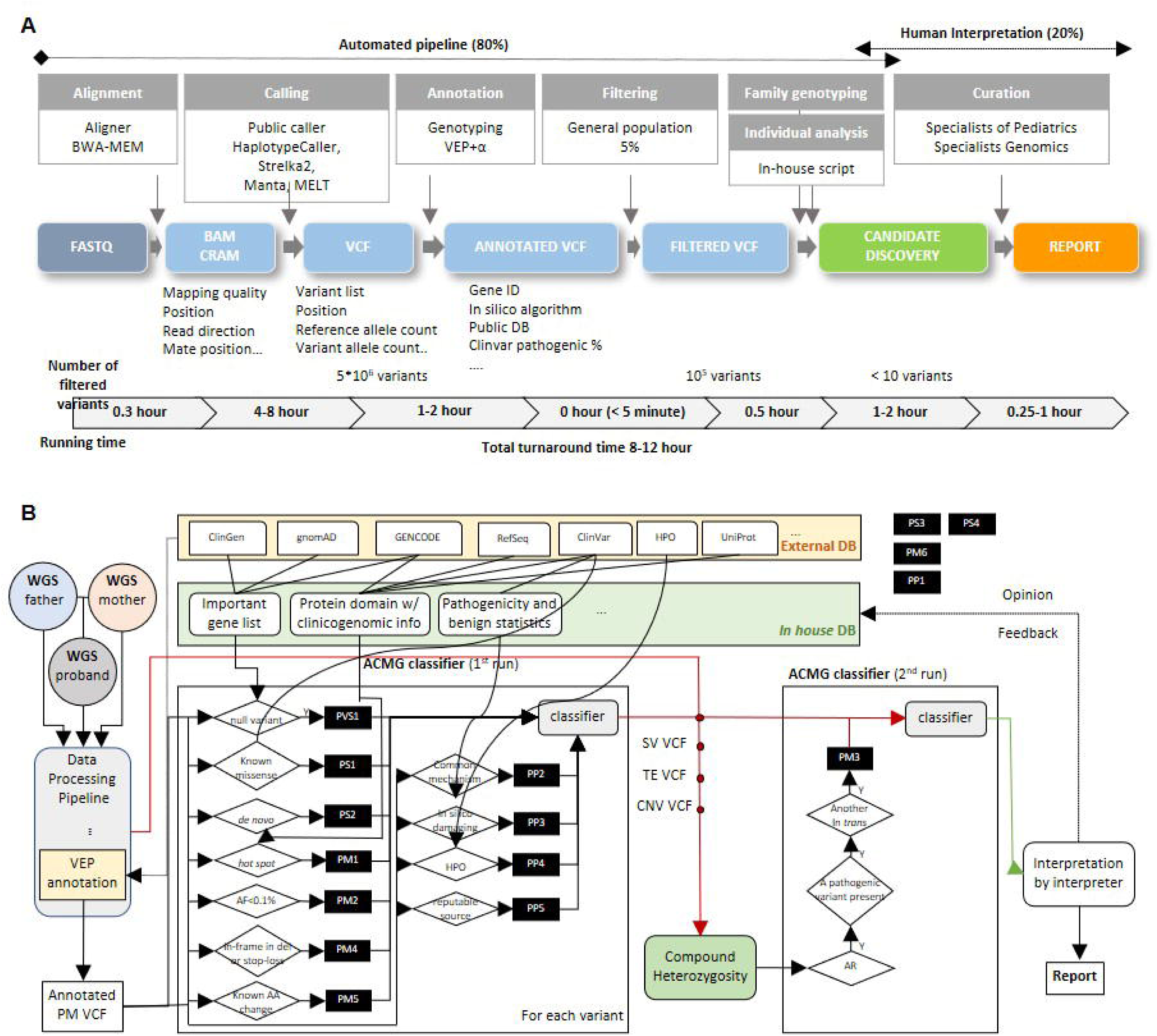
Automated pipeline for whole-genome sequencing (WGS) analysis in patients with genetic diseases. (A) Schematic overview of the design of the automated pipeline for WGS analysis. (B) Flowchart of the *in-house* algorithm used for pathogenicity prediction of each variant.

### Development of a pathogenicity prediction algorithm for genetic variants

For SNVs and INDELs, pathogenicity identification was primarily based on the ACMG guidelines (Figure. 1B). PVS1 was assigned to null variants in genes known to be associated with disease, using Gene-Disease Validity data from ClinGen and a pLI score greater than 0.95. PS1 referred to known missense variants documented in ClinVar. ^18^ PS2 was used for de novo variants, confirmed by genetic parent-child relationships. PM1 applied to hotspot mutations within protein domains, curated in an in-house clinicogenomic database, which integrates information from GENCODE, ^19^ RefSeq, ^20^ ClinVar,^18^ and UniProt. ^21^ PM2 was assigned to variants with a population allele frequency below 0.1%, using the maximum frequency across all ethnic groups. PS3 addressed compound heterozygosity and was applied not only to point mutations but also to SVs, TEs, and CNVs. For genes with an autosomal recessive mode of inheritance, if one pathogenic (P) or likely pathogenic (LP) mutation was already present, the presence of a second P/LP variant in trans was considered causative. PM4 included in-frame indels or stop-loss variants. PM5 was used for variants causing amino acid changes known to be associated with disease. PP2 referred to variants linked to a known common mechanism. For example, if a variant was missense but all known P/LP variants near that position were frameshift mutations, and their number was sufficient, the missense variant was considered benign. PP3 was assigned when in silico analyses provided 1.5 times more damaging evidence than benign evidence. PP4 was based on a gene-phenotype correlation obtained from the Human Phenotype Ontology (HPO) database, with a ‘true’ value assigned if the variant’s associated phenotype matched that of the proband. Lastly, PP5 was given a ‘true’ value if the variant corresponded to a known P/LP variant documented in ClinVar and had been reported in more than one PubMed paper.

In the case of SV, variants that disrupt the structure of the transcript were considered as pathogenic. To assess the potential pathogenic effects of CNV, the effect of gene-copy gains, or loss was assessed based on dosage sensitivity data in the ClinGen guideline. In addition, the presence of absence of heterozygosity (AOH) was utilized as an indicator for variants that could be pathogenic when present in a homozygous state. Furthermore, family analysis was conducted on the patients, sequenced along with their biological parents (trio WGS). This analysis is composed of in-house developed algorithms such as Triomix analysis for uniparental disomy (UPD), ^22^ de novo variant identification, segregation analysis, compound heterozygosity, and double-hit. In the case of isodisomy, it is possible to determine from whom the region was inherited. After that, dozens of candidate causative pathogenic variants from the automated pipeline were manually curated by human experts and discussed in molecular board meetings comprising clinicians and genome scientists.

### Optimization techniques for enhancing germline pipeline efficiency

To enhance the efficiency of the germline pipeline, the WGS analysis pipeline implemented several optimization techniques. Overall, many of analyses utilized multithreading, taking advantage of the high memory and multiple cores available in our computing environment, which significantly reduced the processing time. Additionally, the WGS analysis pipeline performed an asynchronous analysis of the steps that determined the overall efficiency, structuring the pipeline as a workflow system to optimize resource utilization. For sequence variant calling, we split the BAM files to enable parallelization because HaplotypeCaller does not inherently support parallel processing. During segment copy number estimation, the whole genome was divided into smaller regions, and multithreading was employed to analyze each segment in parallel, significantly improving time efficiency. When annotating variants with the Ensembl variant effect predictor (VEP), ^23^ we filtered out non-significant variants to avoid detailed annotation and utilized custom annotations by modifying the database sources, which increased the efficiency compared with the default VEP version. In addition, other modules were integrated to further enhance the efficiency of the tools used by VEP. For family genotyping, we focused on annotating allele distributions for the proband’s germline variants and eliminating redundant variant calls for each parent. In the TrioMix analysis, we used known SNV positions to quickly identify abnormal zygosity regions in the proband and infer the inheritance mechanisms.

## Results

### Validation of the *in-house* pathogenicity prediction algorithm

To validate the sensitivity and specificity of the implemented automatic pipeline, we compared the predicted pathogenicity of variants from the pipeline with the reported pathogenicity in the ClinVar database. In an evaluation of 952,819 variants identified in 5,020 individuals, ^24^ 4,067 variants (39.9%) classified as P/LP by ClinVar were also predicted as P/LP by the pipeline, while 3 variants (0.04%) classified as B/LB by ClinVar were predicted as P/LP by the pipeline (Supplementary Figure 1A). To compare the functional impact of variants classified as P, LP, or variant of uncertain significance (VUS) by the *in-house* pathogenicity prediction algorithm, computational tools such as PolyPhen-2, SIFT, and Provean were implemented (Supplementary Figure 1B-C). ^25–27^ Variants predicted as P or LP were found to significantly impair amino acid function compared to VUS. Moreover, variants predicted as P were shown to cause greater functional impairment than those predicted as LP. Additionally, for variants reported as VUS in ClinVar but classified as P/LP by our algorithm, computational tools revealed significantly higher functional impairment compared to those predicted as VUS by our pipeline. These findings suggest that the *in-house* algorithm can provide additional pathogenicity predictions for VUS previously reported in ClinVar.

### Patient demographics

The prospective application was performed in 20 patients (10 female and 10 male) in the real-world setting. The clinical information of all the patients is described in Table 1. The cohort included 11 neonates who were enrolled from the NICU, and 9 patients aged 1–28 months who were referred from the general ward or PICU. Fourteen patients (70.0%) presented with their first symptoms within one month of birth. The main symptoms were neonatal seizures (n = 5), respiratory insufficiency (n = 3), congenital hypotonia (n = 2), cholestasis and progressive hepatic dysfunction (n = 2), lethargy with hyperammonemia and seizure (n = 1), and cardiopulmonary dysfunction (n = 1). Sixteen patients (16/20, 80%) presented neurologic symptoms as their chief complaint: 7 patients with hypotonia and/or seizures from the NICU, and 9 with profound developmental problems, seizures, or encephalopathy from the ward and PICU. All patients enrolled from PICU or ward displayed severe neurologic symptoms. Enrollment decisions were made within one week of symptom onset for 2 patients born at our hospital; however, for the referred patients, the median enrollment time was 73 days (range, 6–335 days). Two patients had older siblings who displayed similar clinical presentations and died before their genetic etiologies were identified. Clinical outcomes varied among the patients, ranging from death in 2 cases to normal development in 3 cases.

**Table 1.**
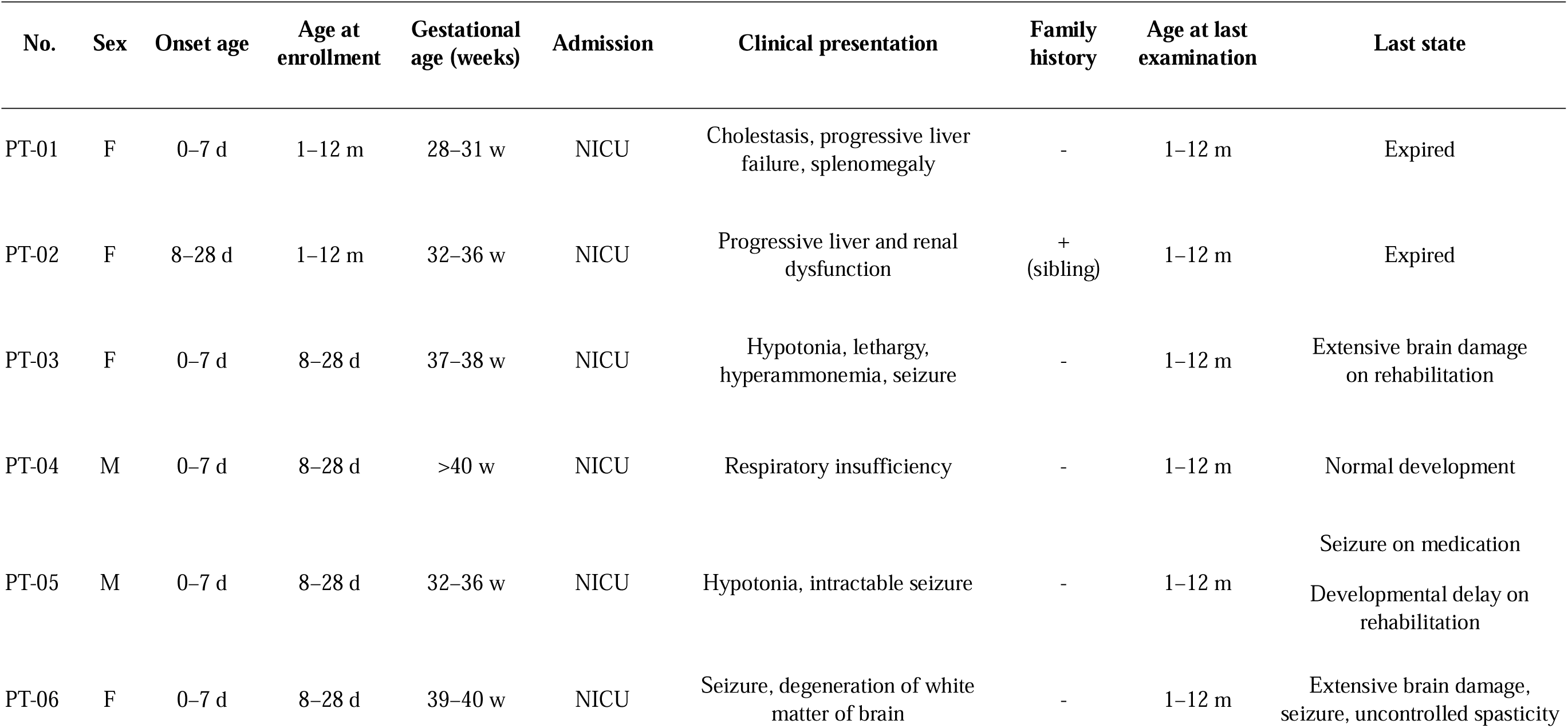
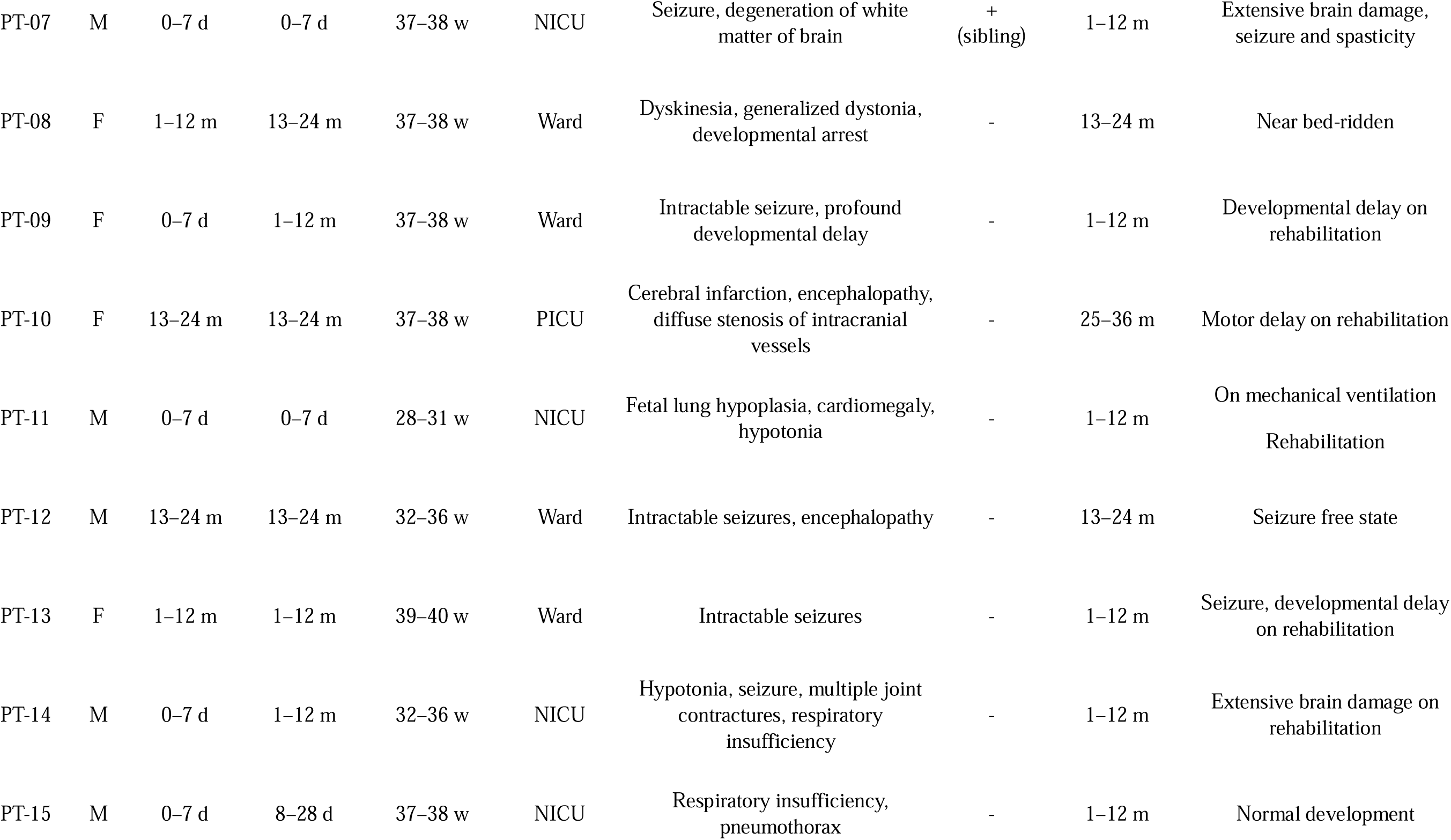
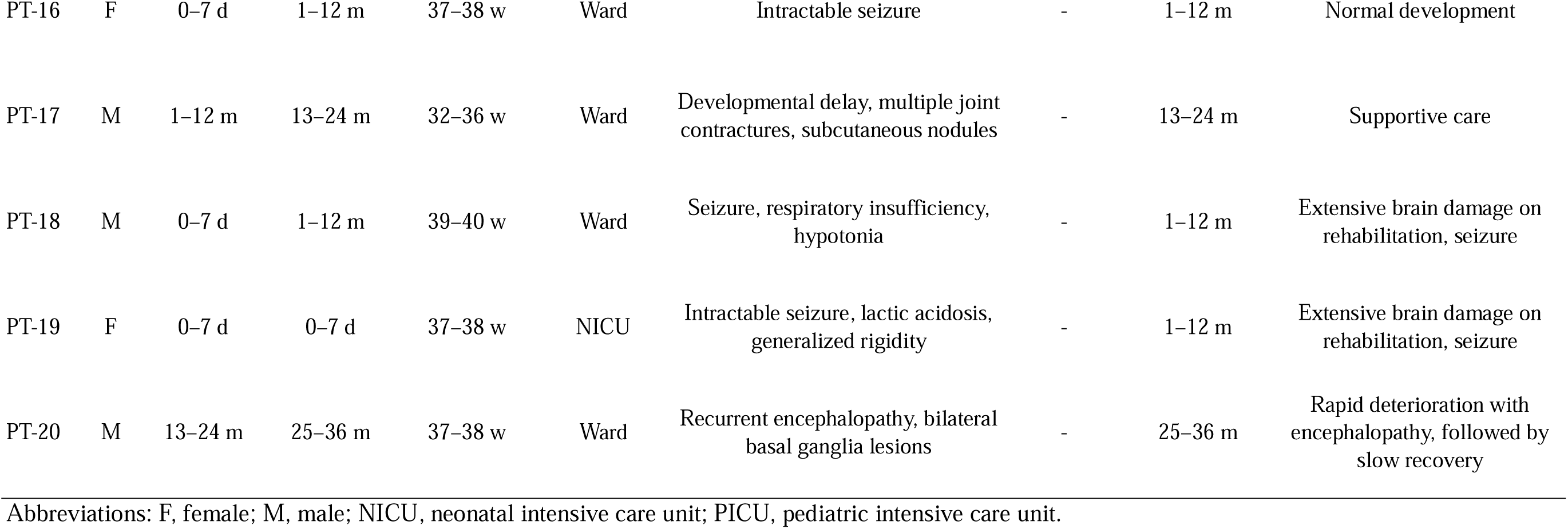
Clinical information of patients.

### Genomic variants detected through rapid whole-genome sequencing

Rapid sequencing was conducted with a median turnaround time (TAT) of 10 days, ranging from 5 to 21 days (Supplementary Figure 2A-B and Supplementary Table 1). A detailed analysis of the time taken for each step revealed that library preparation required a median of 5 days (2–13 days), and sequencing data production took a median of 2 days (0–6 days). Notably, from the production of sequencing data to the final report generation, which involved an automated pipeline and review by a genetic specialist, only a median of six days (2–11 days) was required. The quality of the sequencing data was robust, with a median sequencing depth of 35.37× (24.16–43.46×) and 75.92% (16.83– 89.84%) of the bases were covered at a depth of 30× or greater. None of the samples, including those from the proband and family members, failed the quality control criteria, ensuring that all the samples were successfully analyzed (Supplementary Table 1).

As shown in Table 2, we identified 14 pathogenic or likely pathogenic variants in 10 of the 20 patients (50.0%). Although most of the identified variants were SNVs or indels, PT-16 had a large heterozygous deletion at 16p13.11, along with a *KCNQ2* variant (c.1611del, p.Val538SerfsTer27). In addition, PT-09 had 2 pathogenic variants, *KCNQ2* (c.740C>T, p.Ser247Leu) and *MYT1L* (c.1475_1476del, p.Tyr492TrpfsTer2), both of which were *de novo* and associated with neurodevelopmental disorders. As a result, 2 of the 10 genetically diagnosed patients (20.0%) were shown to have multiple pathogenic variants.

**Table 2.** Variants profile of the positive cases and their clinical implications.

| ID | Gene symbol | Disease | Variant information | Origin /Zygosity | ClinVar | gnomAD (All/EAS) | ACMG class | TAT (days) | Clinical implications |
| --- | --- | --- | --- | --- | --- | --- | --- | --- | --- |
| PT-02 | *IARS1* | Growth retardation, impaired intellectual development, hypotonia, and hepatopathy (MIM#617093) | c.1633_1634del (p.Lys545GlufsTer4) | Father /Het | None | 9.91E-06 /None | LP | 10 | Discontinuation of life-sustaining treatment |
|  |  |  | c.3548C>T (p.Pro1183Leu) | Mother /Het | None | None /None | VUS |  |  |
| PT-03 | *ASS1* | Citrullinemia (MIM#215700) | c.970G>A (p.Gly324Ser) | Father /Het | P, LP | 5.14E-05 /1.11E-04 | P | 8 | Medication change, special formula, decision of liver transplantation, next baby planning via PGT-M, National welfare registry |
|  |  |  | c.421-2A>G | Mother /Het | P | 9.91E-06 /3.34E-04 | P |  |  |
| PT-05 | *SCN2A* | Developmental and epileptic encephalopathy (MIM#613721) | c.5287T>G (p.Ser1763Ala) | De novo /Het | None | None /None | LP | 5 | Medication change |
| PT-06 | *MOCS2* | Molybdenum cofactor deficiency B (MIM#252160) | c.78T>C (p.Ter89Glnext*3) | Both /Hom | LP, VUS | 8.05E-06 /2.45E-04 | LP | 9 | Vitamin B1 supplementation, low-protein diet, next baby planning via PGT-M, National welfare registry, |
| PT-07 | *MOCS1* | Molybdenum cofactor deficiency A (MIM#252150) | c.684_692del (p.Glu228_Leu230del) | Both /Hom | None | 2.48E-06 /8.91E-05 | LP | 10 | Vitamin B1 supplementation, low-protein diet, next baby planning via PGT-M, National welfare registry |
| PT-08 | *GRIN2A* | Epilepsy, focal, with speech disorder and with or without impaired intellectual development (MIM#245570) | c.1957A>G (p.Met653Val) | De novo /Het | LP | None /None | P | 6 | - |
| PT-09 | *KCNQ2* | Developmental and epileptic encephalopathy 7 (MIM#613720) | c.740C>T (p.Ser247Leu) | De novo /Het | P, LP | None /None | P | 8 | - |
|  | *MYT1L* | Intellectual developmental disorder, autosomal dominant 39 (MIM#2616521) | c.1475_1476del (p.Tyr492TrpfsTer2) | De novo /Het | None | None /None | P |  |  |
| PT-11 | *RYR1* | Congenital myopathy 1A, autosomal dominant (MIM#117000) | c.14582G>T (p.Arg4861Leu) | De novo /Het | None | None /None | P | 9 | Consultation to rehabilitation and orthopedics, National welfare registry |
| PT-13 | *CDKL5* | Developmental and epileptic encephalopathy 2 (MIM#300672) | c.476del (p.Asn159IlefsTer69) | De novo /Hemi | None | None /None | P | 21 | National welfare registry |
| PT-16 | *KCNQ2* | Seizures, benign neonatal 1 (MIM#121200) | c.1611del (p.Val538SerfsTer27) | Father /Het | None | None /None | LP | 15 | - |
|  | - | - | 16p13.11 microduplication (chr16:14893103-16512134) | Father /Het | VUS | 1.56E-03 /2.01E-03 | P<sup>a</sup> |  |  |
| PT-17 | *ASAH1* | Farber lipogranulomatosis (MIM#228000) | c.703G>C (p.Gly235Arg) | Father /Het | P | None /None | LP | 10 | Next baby planning via PGT-M, National welfare registry |
|  |  |  | c.256dup (p.Thr86AsnfsTer14) | Mother /Het | P | 6.28E-07 /2.24E-05 | P |  |  |

<sup>a</sup> Predicted by AnnotSV
Abbreviations: EAS, East Asian; TAT, turnaround time; LP, likely pathogenic; VUS, variant of uncertain significance; P, pathogenic; PGT-M, preimplantation genetic testing for monogenic disorder.

Interestingly, in PT-07, we identified a rare homozygous pathogenic variant of the *MOCS1* gene inherited from both parents, with a gnomAD allele frequency of <0.1% (Figure. 2A). Chromosome view analysis, which evaluated read depth and B allele frequency, revealed the AOH not only on chromosome 6 but also across multiple chromosomes (Figure. 2B). To gain a deeper understanding of the multiple AOH regions identified in the patient, we used the TrioMix algorithm^22^ to assess the inheritance patterns of the SNV groups (Figure. 2C). This analysis revealed two distinct inheritance patterns in the patient SNV variant allele fraction (VAF) distribution, strongly suggesting the presence of a common ancestor between the parents (Figure. 2D). SNVs in Group A were identified as shared segments of common ancestral origin inherited from both parents, resulting in a VAF of 1 for heterogeneously altered alleles. Conversely, SNVs in Group B, inherited as shared segments from either parent, had a VAF of 0 for the heterogeneously altered alleles. These findings indicate a limitation in the gene pool and a genetic bottleneck effect across generations, further supporting the hypothesis of a common ancestor between the parents. Due to the possibility of shared pathogenic variants other than the *MOCS1* mutation, we conducted a genome-wide screening in both parents as part of their planning for the next baby, and no additional shared pathogenic variants were identified.

**Figure. 2.**
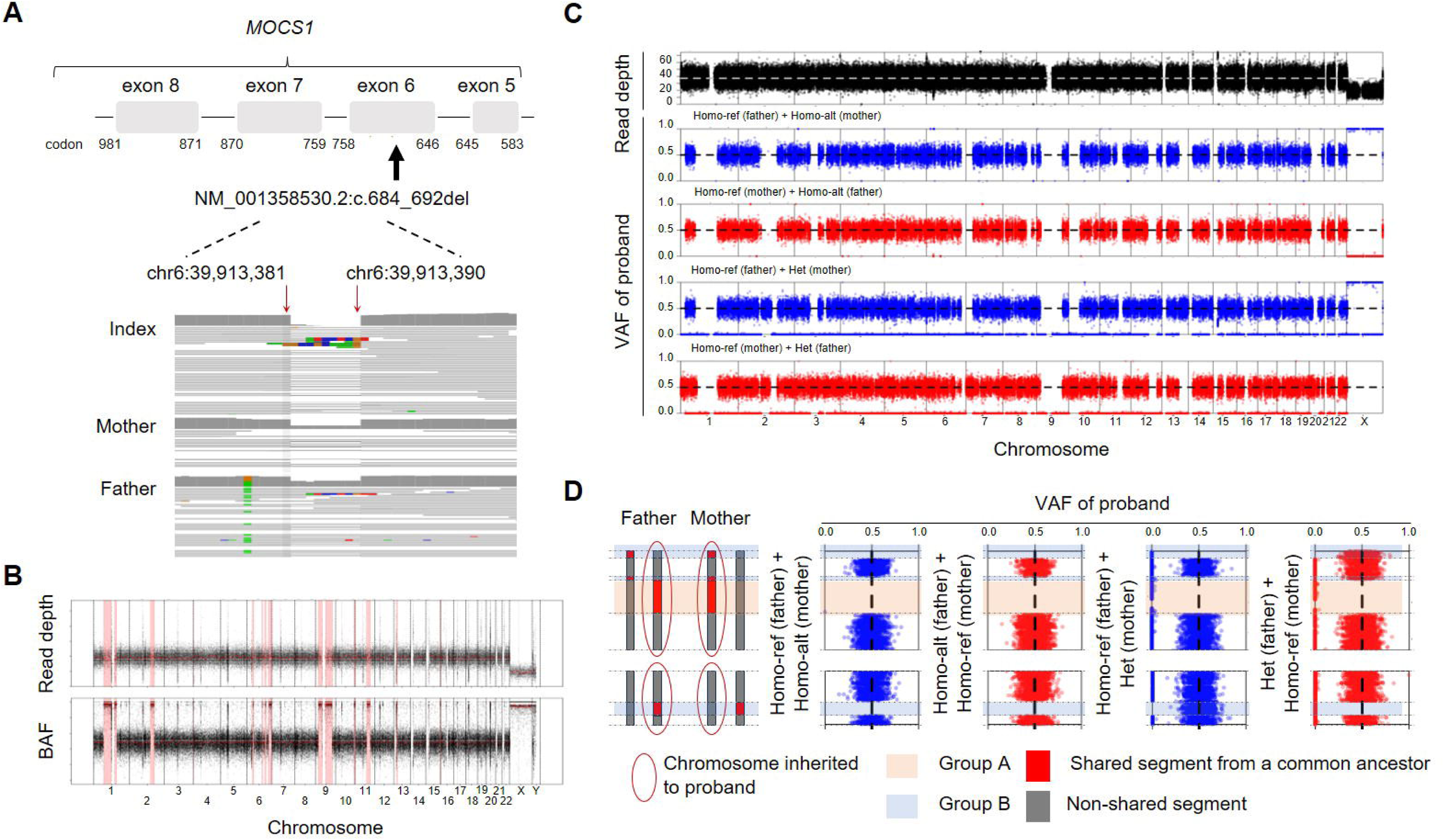
Identification of homogeneous rare pathogenic variants suggesting consanguinity. (A) Schematic illustration showing the location of the identified pathogenic variant in *MOCS1* in the proband (top). The genomic regions corresponding to the variant visualized using the Integrative Genomics Viewer (bottom). (B) Chromosome view showing the average read depth and B allele frequency for binned genomic regions of the proband. (C and D) The TrioMix diagram showing the average read depth for binned genomic regions, the variant allele fraction of the proband’s single nucleotide variants according to inheritance pattern (C), and schematic illustration (D).

In PT-02, we identified an additional VUS and a likely pathogenic variant, both detected in trans in *IARS1*, which causes growth retardation, impaired intellectual development, hypotonia, and hepatopathy (MIM#617093). Although there have been few case reports to date, most reported cases presented with hepatopathy, and some patients died early because of liver failure. ^28, 29^ As PT-02 showed a similar clinical presentation and had an affected sibling who had died earlier, we presumed that *IARS1* was the causative gene. Considering the presumed diagnosis, the diagnostic rate of rWGS in this study was 55.0%.

Through a diagnostic work-up after rWGS, it was suspected that 2 patients developed their condition due to non-genetic causes. PT-12 was diagnosed as a myelin-oligodendrocyte glycoprotein antibody-associated disease using immunoassays and improved after corticosteroid treatment. PT-15 presented with respiratory distress, bilateral lung haziness, and pulmonary hypertension, which were suspected symptoms of surfactant dysfunction. The patient received intensive care with extracorporeal membrane oxygenation, was discharged without any complications, and remained healthy for several months, suggesting a non-genetic condition. When patients with suspected non-genetic etiology were excluded, the diagnostic rate of rWGS was estimated to be 61.1% (11/18 patients).

In this study, 10 of 11 diagnosed patients presented with severe neurological conditions, with identified variants linked to genetic disorders primarily manifesting neurological symptoms. Consequently, the diagnostic rate among the 16 patients with suspected genetic causes and predominant neurological symptoms was 62.5% (10/16 patients), significantly higher than the 25% (1/4) observed in patients with non-neurological symptoms. These findings highlight the particular effectiveness of rWGS in the diagnosis and management of neuro-critical patients.

### Clinical implication of identified pathogenic variants

Management plans were changed in 8 of the 11 (72.7%) genetically diagnosed patients (Table 2). Medication changes and diet modifications were implemented in 4 (36.4%) and 3 (27.3%) patients, respectively. PT-03, referred after undergoing cardiopulmonary resuscitation at another center, had received intravenous L-arginine and continuous renal replacement therapy (CRRT) for hyperammonemia at the time of study enrollment based on the clinical impression of secondary multiorgan failure or a metabolic disorder. The patient was diagnosed with citrullinemia using rWGS and subsequently managed with sodium benzoate, sodium phenylbutyrate, L-carnitine, and a modified diet. Two patients diagnosed with molybdenum cofactor deficiency were started on thiamine supplementation and a low-protein diet. PT-05, who had intractable seizures, was treated with additional phenytoin and carbamazepine, which resulted in a 75% reduction in seizures. The family of PT-02, who presented with progressive hepatopathy and multiorgan dysfunction chose not to pursue invasive treatments, including CRRT. Genetic counseling was provided to all patients and their families, with 4 families (36.4%) planning preimplantation genetic testing for monogenic disorders (PGT-M) for future pregnancies. Six patients (54.5%) were registered with the National Health Insurance Service (NHIS) system in Korea for specific rare genetic disorders and received support for medical expenses.

## Discussion

This study demonstrated the clinical utility of rWGS in critically ill patients in Korea with a diagnostic yield of 55%, which is consistent with previous studies. ^30–33^ Numerous studies on rWGS and rapid whole-exome sequencing, including national project, have focused on critically ill patients. ^33–35^ Technical advances have enabled testing within a few days; however, owing to limited resources, it is necessary to clarify the criteria in most countries. Although the enrollment criteria varied among studies, most included critically ill pediatric patients admitted to the ICU. This study applied similar criteria, but also included patients in the general ward because many patients are transferred from outside NICUs or PICUs to the general ward in tertiary hospitals after acute management in Korea. In this study, all except 2 patients were referred from other hospitals. Among them, 7 patients were enrolled from the general ward and 5 were diagnosed, resulting in a high positivity rate compared with the total study population (71.4% vs. 50.0%). Given that only a few tertiary hospitals provide rWGS, this strategy has the advantage of benefiting a large number of patients.

Although meta-analyses have failed to achieve statistical significance, ^36, 37^ WGS is expected to provide additive diagnostic value compared to gene panels or whole-exome sequencing by detecting various types of variants, such as CNVs and SVs, which might be missed by targeted approaches. In addition, a study reported that the likelihood of a diagnosis is significantly higher for trios than for singletons. ^37^ Although a randomized controlled trial of rapid sequencing did not find a significant difference in diagnostic rates between trio and singleton sequencing, it demonstrated that trio sequencing could aid in achieving a faster TAT. ^38^ Therefore, this study performed rWGS in trios to enable quicker and more accurate diagnoses, thereby achieving a diagnostic rate of >50%. This diagnostic rate is comparable to those of previous studies. ^30–33, 36–39^

Onset age has been another factor in the enrollment criteria for rWGS studies. Most studies have limited their participants to pediatric populations, particularly neonates and infants. ^40^ While some studies included toddlers and adolescents, the proportion of patients aged >3 years ranged from 10–20%,^32, 41, 42^ indicating that the majority of participants were neonates and infants. In this context, this study also included patients aged <3 years old. It is clear that the rWGS can provide clinical benefits to older patients with medical crises caused by genetic etiologies. In previous studies, however, older patients were often diagnosed with non-progressive and incurable genetic disorders. ^32, 41, 42^ Considering the limited resources, different and more detailed criteria for older patients may be a reasonable strategy for real-world practice. Additionally, the diagnostic rate and the proportion of management changes according to primary manifestations can be considered when implementing an rWGS platform. In this study, the diagnostic yield was quite higher in patients with neurological manifestations (62.5%, 10/16), compared to those with non-neurological manifestations (25%, 1/4). The small sample size made it difficult to draw precise conclusions, but the findings suggested that rWGS is more likely to provide valuable information for patients with severe neurological symptoms. Further studies could help establish clearer criteria in resource-limited setting.

The clinical utility of this study is evident. Clinicians changed or added management plans in 8 of 11 patients with a final or presumed diagnosis (72.7%), which is favorable compared to previous studies. ^31, 40^ Selecting the appropriate antiseizure medication is crucial for patients with early onset epileptic encephalopathy because refractory seizures can cause secondary brain damage and further developmental decline. Specific dietary modifications and vitamin supplementation are often the only options for the treatment of certain metabolic disorders. As most participants were neonates and infants, counseling for future family planning is a critical and urgent issue. In our cohort, 5 genetic disorders with recessive inheritance patterns were identified, including two patients with siblings who died early due to a similar presentation (PT-02 and PT-07). Four families began preparing PGT-Ms for subsequent pregnancies. In particular, for PT-07, whose parents were suspected to have a common ancestor, we further evaluated other pathogenic or likely pathogenic variants shared by the mother and father before PGT-M. These findings highlight the clinical utility of genome-wide screening facilitated by rWGS.

Several studies have documented the economic benefits of early diagnosis. ^7, 37, 40^ In this study, a systematic cost-effectiveness analysis was not performed because of the small study population and the varied clinical settings. Nonetheless, the total medical cost has decreased owing to shorter hospitalizations and the avoidance of unnecessary tests and treatments. In addition, timely diagnosis is invaluable for critically ill patients and their families. Government support through the National Disease Registry also has an important impact on the family, as the registry can often be completed after genetic diagnosis in Korea. In this study, more than half of the diagnosed patients (6/11, 54.5%) were registered with the NHIS in Korea through a genetic diagnosis.

Effective patient enrollment and shortening of the TAT are issues that need to be addressed before implementing rWGS in Korea. Although more than 100 regional hospitals operate NICUs (https://www.hira.or.kr/), most do not have the capability for rWGS. All except two patients in this study were initially treated at other hospitals, leading to delays in rWGS, as enrollment could only be performed after referral. The average time from symptom onset to testing was 72 days in the referred group, compared to 4 and 7 days for the two patients directly admitted to the study hospital. To achieve maximum performance with the rWGS platform, a fast-track patient referral system or consignment test may be necessary in real-world practice.

The TAT is another crucial factor for rWGS platforms in critical care. Efforts to reduce TAT are ongoing, with recent studies reporting TAT hours from sample collection to reporting. ^41, 43, 44^ In this study, the average TAT was approximately 10 days, ranging from 5 to 21 days, which is relatively long compared with previous reports. The primary factors contributing to delayed reporting are processes that require human intervention, such as shipping, library preparation, and report generation. Once library preparation was completed, the production of sequencing data and subsequent pipeline analyses were predominantly automated, allowing the completion of these processes in a relatively short and stable timeframe. Therefore, to further reduce the TAT, systematizing and minimizing the need for human intervention at these stages could be highly effective. Automating these human resource-intensive processes can streamline workflow, significantly decrease the overall TAT, and improve the efficiency and responsiveness of rWGS platforms in critical care environments.

### Conclusion

This study demonstrates the feasibility and clinical utility of rWGS in Korea. rWGS enables timely management and family counseling for critically ill pediatric patients, particularly among those presenting with neurologic symptoms. Establishing detailed testing indications and a nationwide fast-track testing system may be necessary for its effective implementation in clinical practice.

## Author contribution

Conceptualization: Kim SY, Chae JH. Methodology: Lee S, Koh JY, Lim J, Lee Y, Yi BR, Joo EJ, Jung D. Data acquisition: Lee S, Cho J, Lim BC. Analysis: Koh JY, Lim J, Kim W, Lee Y, Yi B, Jung D, Kim SY. Interpretation: Lee S, Koh JY, Lim J, Cho J, Kim W, Lee Y, Yi B, Joo E, Jung D, Limb BC, Kim SY. Investigation: Lee S, Koh JY, Lim J, Cho J, Kim W, Lim BC, Kim SY. Visualization: Koh JY, Lim J. Funding acquisition: Chae JH, Lee S. Project administration: Kim SY, Lee S, Koh JY. Supervision: Chae JH. Writing – original draft: Kim SY, Lee S, Koh JY, Lim J. Writing – review & editing: Kim SY, Chae JH.

## Statements and Declarations

### Ethical consideration

The study protocol was approved by the Institutional Review Board of Seoul National University Hospital (approval no. 2303-168-1417), and compliance with the relevant guidelines and regulations was ensured.

## Consent to participate

Informed consent was obtained from the parents or legal guardians of all participants prior to blood sample collection.

## Consent for publication

Not applicable

## Data availability

The datasets generated and analysed during the current study are not publicly available but are available from the corresponding author on reasonable request.

