## Supplement data for "Trio-Based Whole-Genome Sequencing for Critically Ill Pediatric Patients in Korea"

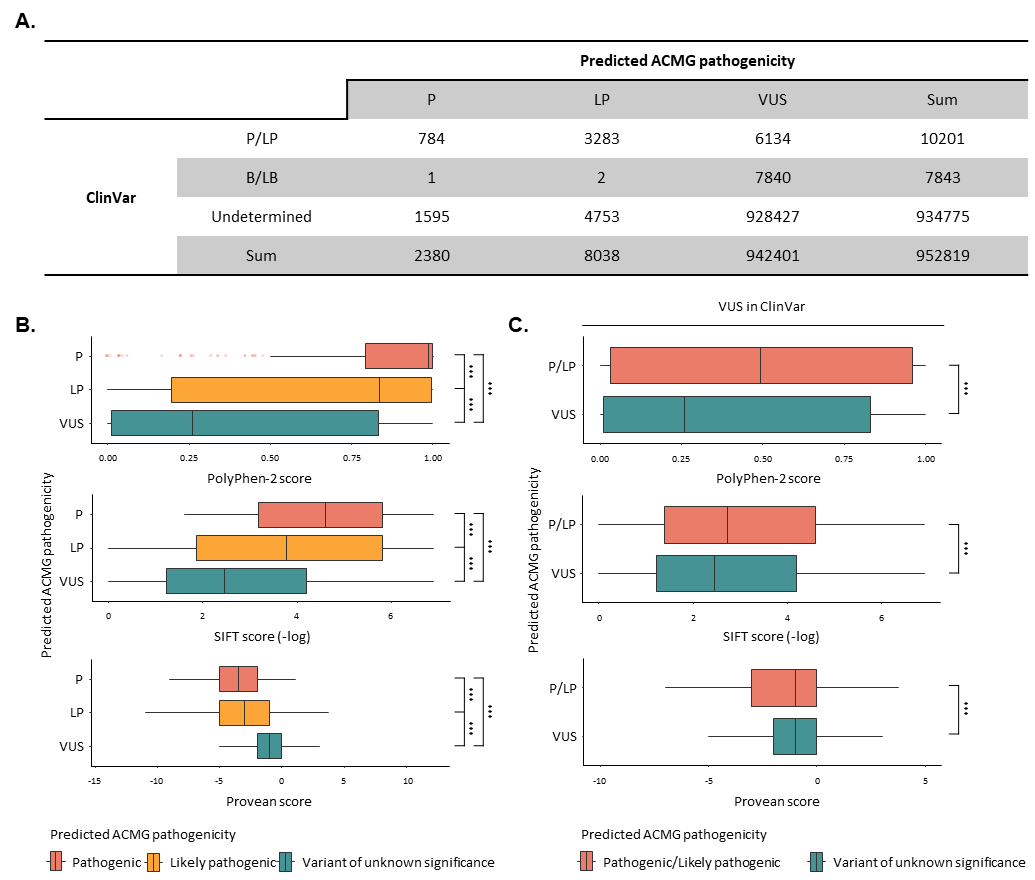

**Supplementary Figure 1. Validation of the *in-house* pathogenicity prediction algorithm.** (A) Table comparing the pathogenicity classification of each variant as reported by the ClinVar database and as predicted by the *in-house* pathogenicity prediction algorithm. (B) Bar plots showing functional impact scores of variants classified as pathogenic (P), likely pathogenic (LP), or variant of uncertain significance (VUS) by the *in-house* algorithm, using computational tools, including PolyPhen-2, SIFT, and Provean. (C) Bar plots showing functional impact scores for variants classified as P/LP or VUS by the *in-house* algorithm but reported as VUS in ClinVar.

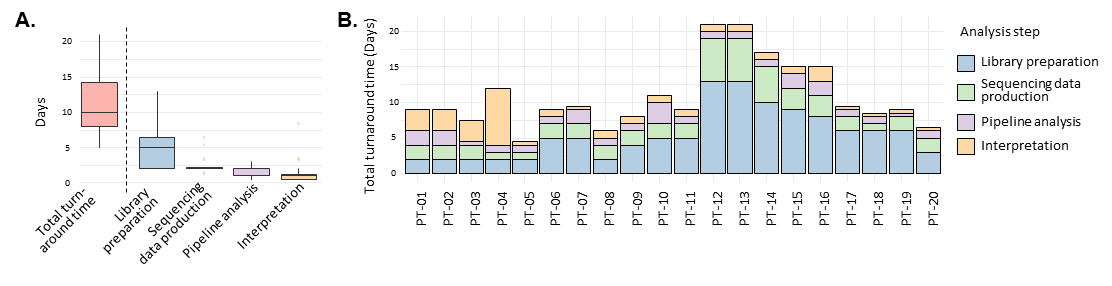

**Supplementary Figure 2. Running time for the automated pipeline in whole-genome sequencing (WGS) analysis.** (A-B) Bar plots showing the running time for each step of the pipeline, presented for the entire cohort (A) and for individual samples (B).

**Supplementary Table 1. Metrics for sequencing process.**

| **No** | **Turnaround time (days)** | | | | |  | **QC metrics** | | |
| --- | --- | --- | --- | --- | --- | --- | --- | --- | --- |
|  | **Total** | **Library preparation** | **Sequencing data production** | **Pipeline analysis** | **Report generation** |  | **Mean read depth** | **Percent of duplicate read (%)** | **% Covered genomic regions (≥30x)**^†^ |
| PT-01 | 10 | 2 | 2 | 2 | 3 |  | 35.92 | 11.07 | 82.86 |
| PT-02 | 10 | 2 | 2 | 2 | 3 |  | 31.92 | 10.74 | 64.32 |
| PT-03 | 8 | 2 | 2 | <1 | 3 |  | 25.52 | 7.57 | 24.24 |
| PT-04 | 14 | 2 | 1 | 1 | 8 |  | 42.54 | 7.66 | 89.84 |
| PT-05 | 5 | 2 | 1 | 1 | <1 |  | 41.74 | 7.36 | 89.09 |
| PT-06 | 9 | 5 | 2 | 1 | 1 |  | 30.87 | 8.18 | 58.47 |
| PT-07 | 10 | 5 | 2 | 2 | <1 |  | 37.68 | 9.46 | 85.11 |
| PT-08 | 6 | 2 | 2 | 1 | 1 |  | 27.71 | 7.89 | 39.2 |
| PT-09 | 8 | 4 | 2 | 1 | 1 |  | 27.5 | 9.68 | 36.6 |
| PT-10 | 11 | 5 | 2 | 3 | 1 |  | 25.46 | 11.37 | 24.17 |
| PT-11 | 9 | 5 | 2 | 1 | 1 |  | 40.55 | 1.24 | 80.87 |
| PT-12 | 21 | 13 | 6 | 1 | 1 |  | 39.88 | 1.03 | 81.9 |
| PT-13 | 21 | 13 | 6 | 1 | 1 |  | 37.94 | 0.7 | 82.95 |
| PT-14 | 17 | 10 | 5 | 1 | 1 |  | 39.47 | 0.97 | 81.94 |
| PT-15 | 16 | 9 | 3 | 2 | 2 |  | 39.36 | 0.88 | 81.27 |
| PT-16 | 15 | 8 | 3 | 2 | 2 |  | 39.57 | 0.79 | 84.6 |
| PT-17 | 10 | 6 | 2 | 1 | <1 |  | 35.37 | 1.04 | 74.63 |
| PT-18 | 8 | 6 | <1 | 1 | <1 |  | 29.9 | 0.6 | 56.45 |
| PT-19 | 8 | 6 | 2 | <1 | <1 |  | 33.16 | 0.79 | 69.55 |
| PT-20 | 6 | 3 | 2 | 1 | <1 |  | 27.72 | 0.57 | 46.5 |

^†^ The percentage of bases covered at a depth of 30x or greater.
